# Demographic characteristics of SARS-CoV-2 B.1.617.2 (Delta) variant infections in Indian population

**DOI:** 10.1101/2021.09.23.21263948

**Authors:** Ashutosh Kumar, Adil Asghar, Khursheed Raza, Ravi K. Narayan, Rakesh K. Jha, Abhigyan Satyam, Gopichand Kumar, Prakhar Dwivedi, Chetan Sahni, Chiman Kumari, Maheswari Kulandhasamy, Rohini Motwani, Gurjot Kaur, Hare Krishna, Kishore Sesham, Sada N. Pandey, Rakesh Parashar, Kamla Kant, Sujeet Kumar

## Abstract

**Importance:** Higher risks of contracting infection, developing severe illness and mortality are known facts in aged and male sex if exposed to the wild type SARS-CoV-2 strains (Wuhan and B.1 strains). Now, accumulating evidence suggests greater involvement of lower age and narrowing the age and sex based differences for the severity of symptoms in infections with emerging SARS-CoV-2 variants. Delta variant (B.1.617.2) is now a globally dominant SARS-CoV-2 strain, however, current evidence on demographic characteristics for this variant are limited. Recently, delta variant caused a devastating second wave of COVID-19 in India. We performed a demographic characterization of COVID-19 cases in Indian population diagnosed with SARS-CoV-2 genomic sequencing for delta variant.

**Objective:** To determine demographic characteristics of delta variant in terms of age and sex, severity of the illness and mortality rate, and post-vaccination infections.

**Design:** A cross sectional study

**Setting:** Demographic characteristics, including vaccination status (for two complete doses) and severity of the illness and mortality rate, of COVID-19 cases caused by wild type strain (B.1) and delta variant (B.1.617.2) of SARS-CoV-2 in Indian population were studied.

**Participants:** COVID-19 cases for which SARS-CoV-2 genomic sequencing was performed and complete demographic details (age, sex, and location) were available, were included.

**Exposures:** SARS-CoV-2 infection with Delta (B.1.617.2) variant and wild type (B.1) strain.

**Main Outcomes and Measures:** The patient metadata containing details for demographic and vaccination status (two complete doses) of the COVID-19 patients with confirmed delta variant and WT (B.1) infections were analyzed [total number of cases (N) =9500, N_delta_=6238, N_WT_=3262]. Further, severity of the illness and mortality were assessed in subsets of patients. Final data were tabulated and statistically analyzed to determine age and sex based differences in chances of getting infection and the severity of illness, and post-vaccination infections were compared between wild type and delta variant strains. Graphs were plotted to visualize the trends.

**Results:** With delta variant, in comparison to wild type (B.1) strain, higher proportion of lower age groups, particularly <20 year (0-9 year: 4.47% vs. 2.3%, 10-19 year: 9% vs. 7%) were affected. The proportion of women contracting infection were increased (41% vs. 36%). The higher proportion of total young (0-19 year, 10% vs. 4%) (p=.017) population and young (14% vs. 3%) as well as adult (20-59 year, 75% vs. 55%) women developed symptoms/hospitalized with delta variant in comparison to B.1 infection (p< .00001). The mean age of contracting infection [Delta, men=37.9 (±17.2) year, women=36.6 (±17.6) year; B.1, men=39.6 (±16.9) year and women= 40.1 (±17.4) year (p< .001)] as well as developing symptoms/hospitalization [Delta, men=39.6(± 17.4) year, women=35.6 (±16.9) year; B.1, men=47(±18) year and women= 49.5(±20.9) year (p< .001)] was considerably lower. The total mortality was about 1.8 times higher (13% vs. 7%). Risk of death increased irrespective of the sex (Odds ratio: 3.034, 95% Confidence Interval: 1.7-5.2, p<0.001), however, increased proportion of women (32% vs. 25%) were died. Further, multiple incidences of delta infections were noted following complete vaccination.

**Conclusions and Relevance:** The increased involvement of young (0-19 year) and women, lower mean age for contracting infection and symptomatic illness/hospitalization, higher mortality, and frequent incidences of post-vaccination infections with delta variant compared to wild type strain raises significant epidemiological concerns.

**Key Points:** *Question:* Did SARS-CoV-2 B.1.617.2 (Delta) variant infections show varied demographic characteristics in comparison to wild type strains?

*Findings:* In this cross sectional study viral genomic sequences of 9500 COVID-19 patients were analyzed. As the key findings, increased involvement of young (0-19 year) and women, lower mean age for contracting infection and symptomatic illness/hospitalization, higher mortality, and frequent incidences of post-vaccination infections with delta variant in comparison to wild type (WT) strain (B.1) were observed.

*Meaning:* The findings of this study suggest that delta variant has varied demographic characteristics reflecting increased involvement of the young and women, and increased lethality in comparison to wild type strains.

## Introduction

Delta variant of SARS-CoV-2 (B.1.617.2) has caused recent COVID-19 waves and spikes in multiple countries and it is now a globally dominant strain^1^. Structural and functional analyses of the lineage characterizing mutations in the spike glycoprotein have predicted potential alterations in virus-host interactions and masking of the antibody binding sites leading to increased transmissibility, lethality, and immune escape capabilities for this variant^2–4^, which were further validated in recent animal model^5,6^ and human studies^7–10^. Available studies precisely indicate that delta is at least 50-60% more transmissible than alpha variant (B.1.1.7)^11^ and are capable of immune escape against the natural infections with previous SARS-CoV-2 strains, the COVID-19 vaccines, and therapeutically used monoclonal antibodies^5,11^.

The emerging SARS-CoV-2 variants have been reported to vary from wild type (WT) strains (Wuhan strain and B.1) in demographic characteristics, such as age and sex based vulnerability for contracting infection, developing severe illness, and risk of mortality^4,12–15^. Against the established pattern of higher vulnerability for the aged and male sex, which is explained by the established immunological reasons^16–20^, the emerging variants are involving young and female sex in increasing proportion^4,12–15^. However, the studies, which have examined the shift in demographic characteristics for the delta variant, are currently limited. Recently, a devastating second COVID-19 wave driven by delta variant occurred in India^11,21^. First COVID-19 wave, similar to other parts of the world, was driven by WT strains in India. First variant with significantly increased transmissibility and lethality was alpha variant, which turned to be a dominant strain by 2020^21^. Alpha variant caused the frequent increase in the daily cases across the Indian states, in between multiple other variants were also reported, however, none of them could lead a significant COVID-19 wave, until the arrival of delta variant^21^. The first case of delta variant was reported from India by December, 2020^21^; however, the second COVID-19 wave was not evident before April, 2021. The second wave continued for months, only seeing a fall in July, 2021^21^.

The demographic characterization of the COVID-19 infections with delta variant across the population groups is an urgent requirement to decide relevant health policy measures preventing recurrent COVID-19 waves. In this study, we analyzed the demographic and associated clinical metadata of COVID-19 cases with SARS-CoV-2 WT (B.1) and delta variant strains in the Indian population, to fulfill that lacunae.

## Materials and methods

### Data collection and processing

(Design, Setting, Participants, Exposure, Data sources/ measurement, Bias, Study size)

A cross-sectional study was conducted for determining the demographic characterization of the SARS-CoV-2 B.1.617.2 (Delta) variant infections in Indian population. The SARS-CoV-2 genomic sequence reports from India (patient sample collection date not later than 31^st^ July, 2021) were accessed from EpiCoV™ database of Global Initiative on Sharing All Influenza Data (GISAID) (https://www.gisaid.org/) using automatic search function feeding information for geographical location, SARS-CoV-2 lineage and collection dates. No further distinction of specific geographical locations within India was made while accessing the data. A similar search strategy was applied to retrieve a comparative metadata for the COVID-19 cases with WT B.1 strain (Wuhan strain with D614G mutation) reported on GISAID since first COVID wave. The metadata files of the delta variant (test group) and WT (control group) strains sequences were downloaded from the individual GISAID accession proportions, checked and confirmed for the accuracy of lineage information, and were screened for the demographic details (age and sex) and vaccination status (for two complete doses). Further, the clinical data (informing severity of the illness, and mortality) of the cases for the WT and delta variant strains were assessed using a repeat search feeding ‘patient status’ as the additional input. The filtering of the collected data was done by discarding the sequence reports containing no or incomplete information, and duplication (repeat sequencing from same individuals). Any selection bias in data collection was taken care of by random sampling for the confounding factors (age, sex, date of collection, and geographical location). Sampling and data entry errors were rechecked and verified.

### Data analysis and presentation

(Quantitative variables, statistical methods)

The final data were spread in the excel sheets and analyzed for the distribution by age and sex. Age distribution of the delta variant infections was presented at ten-year interval. Additionally, for the analysis of clinical outcomes total cases were divided in three broad age groups as follows: ‘young’ (0-19 year), ‘adult’ (20-59 year), and ‘elderly’ (60 year and above). Clinical outcomes of the positive cases were categorized as ‘asymptomatic/mildly symptomatic’ (including the cases in home isolation and/or quarantine with no overt symptoms or mild symptoms), ‘symptomatic/hospitalized’ (including the cases with overt symptoms, currently hospitalized or released/recovered after hospitalization), and ‘demised’. Individual categories were analyzed for the age and sex distribution. For the determination of mortality, the statuses of all the categories other than ‘demise’ were considered as ‘living’. Categorical data were presented as frequency and/or proportions. The continuous data were presented as mean (or median) ± standard deviation (SD).

The statistical tests were performed to evaluate inter-group differences with the help of MS Excel 2019 and XLSTAT package. Normality of the data was examined by Kolmogorov-Smirnov test. Two sample Student’s t test and ANOVA were used for normally distributed data. Games-Howell Post-Hoc Test was applied for inter-group comparisons. Kruskal Wallis H test was used for skewed data. Chi square test was used for categorical variables. Multinomial logistic regression was used for estimation of odd ratio in reference of delta variant versus WT strain. Results were considered statistically significant at p-value ≤ 0.05. Graphs were plotted to visualize the data trends.

## Results

We assessed the genomic sequence reports for a total of 8269 and 3767 cases of delta variant and WT B.1 strains, respectively. After filtering the data, a total of 6238 cases of delta variant and 3262 cases of WT B.1 strain were available for demographic analysis. Among the screened cases 24 incidences of delta infection following two complete doses of vaccines were found, however, no post-vaccination infections were noted with B.1. Similarly, a total of 659 and 320 sequence reports with information on ‘patient status’ were present in the database for delta and B.1 strains respectively, and after filtering of the data 647 and 276 cases were available for analysis of the severity of illness and mortality in terms of age and sex.

### Demographic distribution

The frequent incidences of delta variant as well as B.1 strain infections were noted across the age groups (N=6238), including the young (0-19 year) (Fig. 1). The prevalence of infections was higher with delta variant in comparison to the B.1 strain in young population, the highest in 0-9 year followed by 10-19 year. Conversely, relatively lower infections were noted in > 60 year age groups with fewer exceptions (Fig. 1). Higher proportion of men were infected compared to women with delta variant (59% vs. 41%, N=6238) as well as B.1 (64% vs. 36%, N=3262). The mean age of contracting infection was 37.9 (±17.2) year for men and 36.6 (±17.6) year for women in delta variant infections in comparison to 39.6 (±16.9) and 40.1 (±17.4) years, respectively, in B.1 strain infections (p< .001).

**Figure 1.**
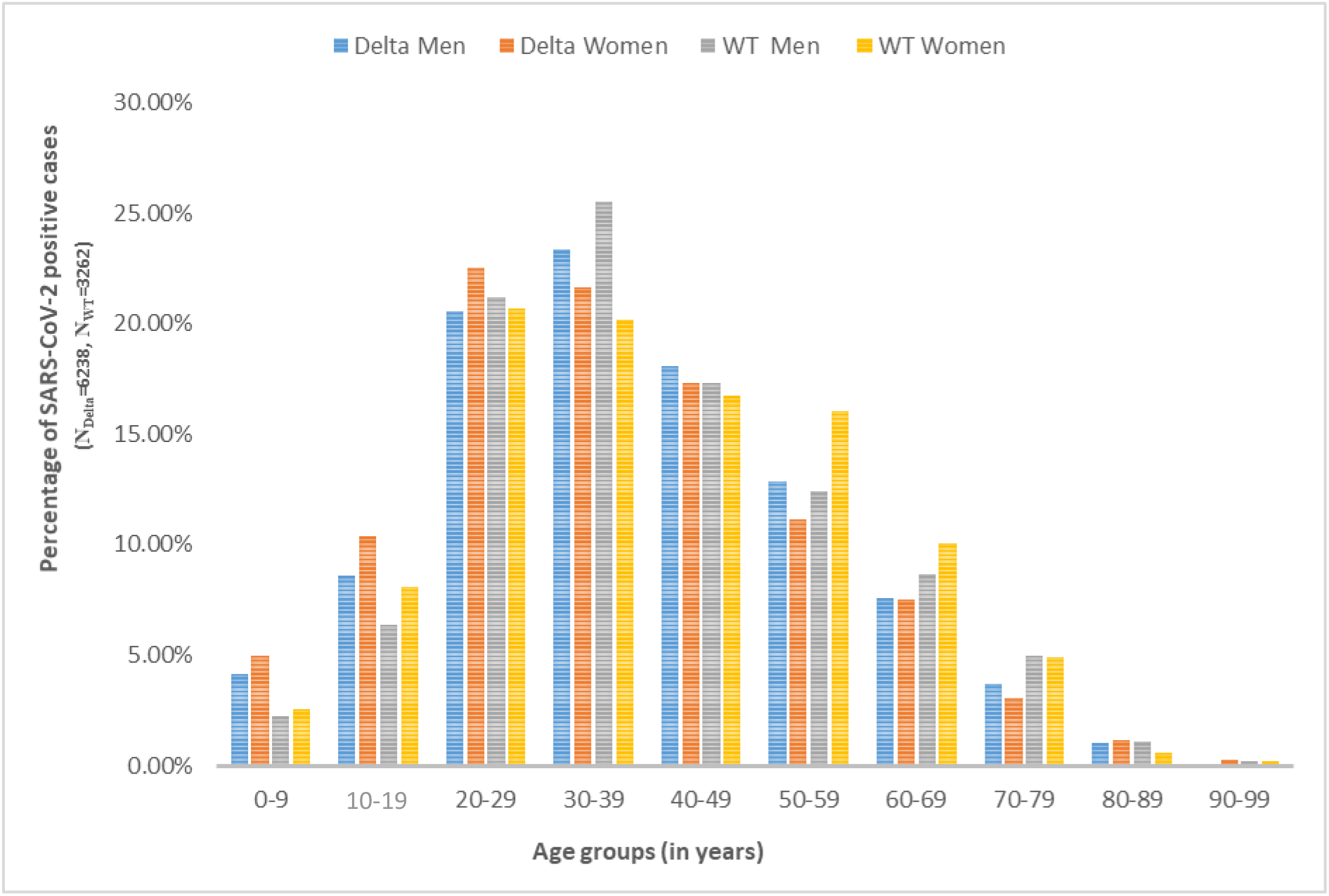
Demographic distribution of SARS-CoV-2 B.1.617.2 (Delta) variant and Wild type (WT) strain (B.1) infections in Indian population.

### Patient status

The cases were noted under all categories for delta variant as well as B.1 (asymptomatic/mildly symptomatic, NDelta =72, NWT=24; symptomatic/hospitalized NDelta =262, NWT=130; living but symptom status not known NDelta =228, NWT=102; and demised, NDelta =85, NWT=20). The demographic distribution of the patient status data in reference to age groups and sex has been shown in Fig. S1.

#### Symptomatic illness and hospitalization

The cases which developed symptoms and required hospitalization were reported in each age group, including the young (0-19 year) for both the strains (Fig. 2a) However, a higher proportion of total symptomatic/hospitalized cases by delta variant were contributed by ‘young (0-19 year) (10% vs. 4%) and adult (75% vs. 66%) in comparison to B.1 (p=.017). Further, a higher proportion of young (14% vs. 3%) and adult (75% vs. 55%) women, albeit not men, developed symptoms and required hospitalization with delta variant in comparison to B.1 infection (p< .00001) (Fig. 2b). The mean age of developing symptoms/hospitalization for men and women was 39.6(± 17.4) and 35.6 (±16.9) years in cases of delta and 47(±18) and 49.5(±20.9) years in cases of B.1, respectively (p< .001).

**Figure 2a.**
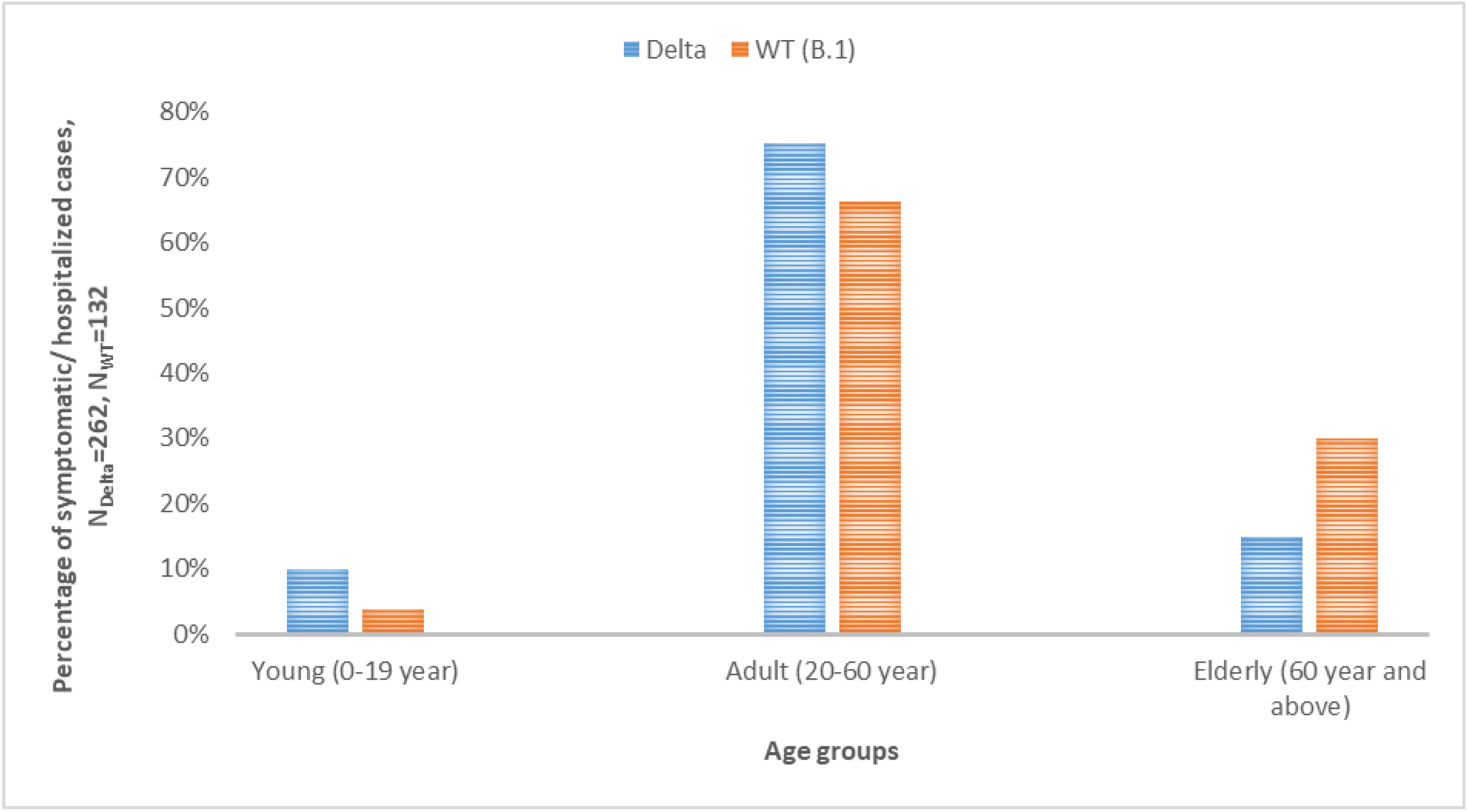
Distribution of ‘symptomatic/hospitalized’ cases with SARS-CoV-2 B.1.617.2 (Delta) variant and Wild type (WT) strain (B.1) infections in reference to age.

**Figure 2b.**
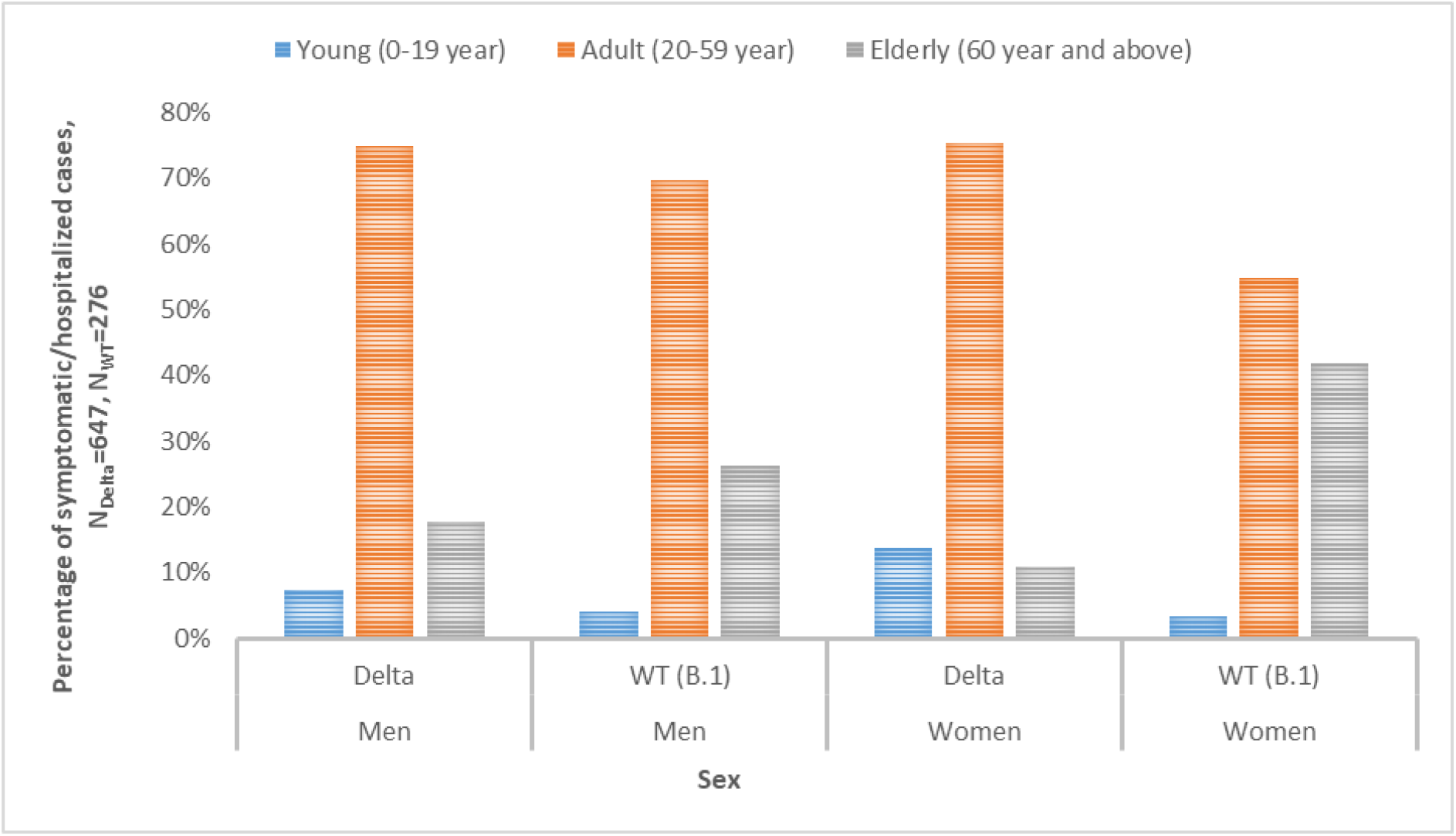
Distribution of ‘symptomatic/hospitalized’ cases with SARS-CoV-2 B.1.617.2 (Delta) variant and Wild type (WT) strain (B.1) infections in reference to sex.

#### Total mortality and risk of death

Out of the total cases, approximately 13 % (85/647) and 7% (20/276) died due to illness by delta and B.1 strains respectively. However, no deaths were reported in the age groups < 20 year for delta as well as B.1 (Fig. 3a). Further, a higher proportion of total mortality was contributed by adult (59% vs. 40%) with delta variant infection in comparison to B.1. Higher proportion of men than women were died by delta variant (68% vs.32%) as well as B.1 (75% vs. 25%). Notably, the proportion of total mortality in women was increased (32% vs. 25%), however, a higher proportion of adult (20-59 year) men (60% vs. 33%) but not women were died with delta variant compared to B.1 (p< .001) (Fig. 3b). The mean age of mortality due to illness for men and women was 56.6(± 13.5) and 58.8(±11.4) years in cases of delta and 60.7(±15.5) and 50.2(±15.7) years in cases of B.1, respectively (p< .001). Risk of death was higher with delta variant infections irrespective of the sex (Odds ratio: 3.034, 95% Confidence Interval: 1.7-5.2, p<0.001). Age but not the sex, was able to predict risk of death. Lower age was marginally protective (Odds ratio: 0.934, 95% Confidence Interval: 0.92-0.95, p<0.001) for both of the strains (Table 1).

**Table 1.** Viral and host factors predicting risk of death.

| Predictor | Z<br>distribution | Statistical<br>significance (p<br>value) | Odds<br>ratio | 95% Confidence<br>Interval |  |
| --- | --- | --- | --- | --- | --- |
|  |  |  |  | Lower | Upper |
| **SARS-CoV-2 lineage:**<br>**B.1-Delta** | 3.93 | < .001 | 3.034 | 1.744 | 5.277 |
| **Age** | -9.10 | < .001 | 0.934 | 0.920 | 0.947 |
| **Sex: Men-women** | -1.11 | 0.267 | 0.765 | 0.477 | 1.227 |

**Figure 3a.**
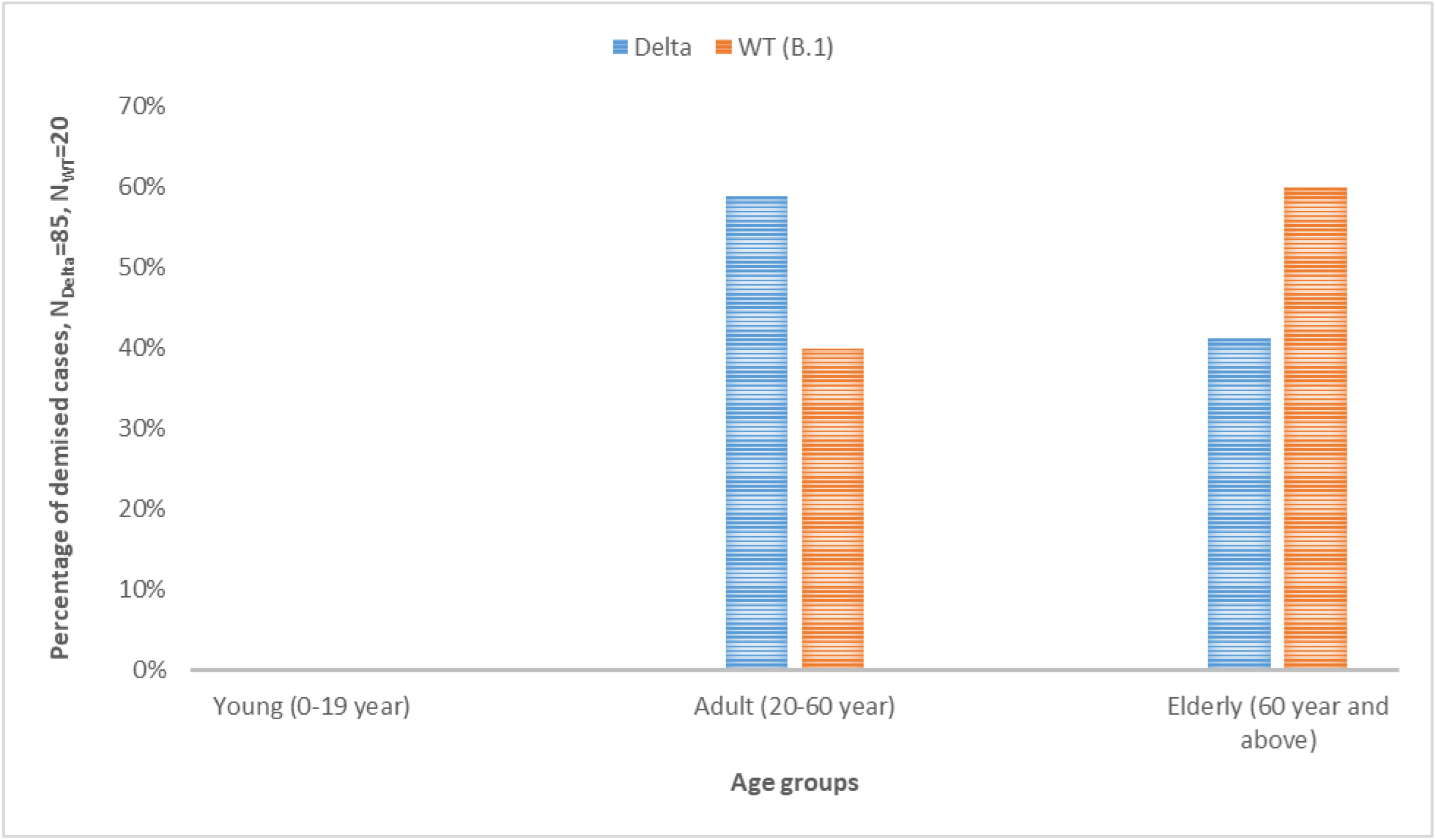
Distribution of ‘demised’ cases with SARS-CoV-2 B.1.617.2 (Delta) variant and Wild type strain (B.1) infections in reference to age.

**Figure 3b.**
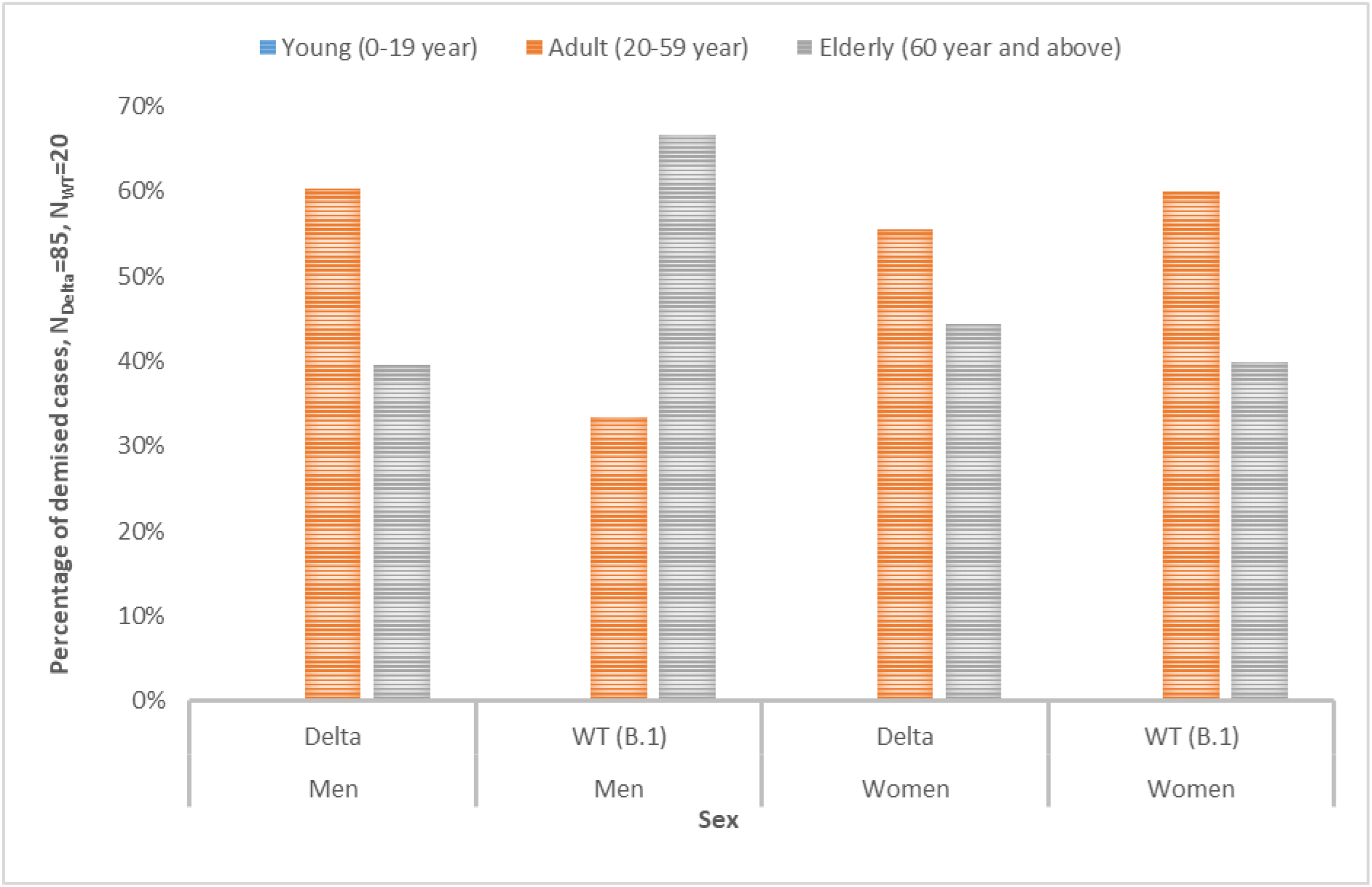
Distribution of ‘demised’ cases with SARS-CoV-2 B.1.617.2 (Delta) variant and Wild type strain (B.1) infections in reference of sex.

#### Post-vaccination infections

We noted the incidences of delta infection following two complete doses of vaccines (N=24, men=12, women=12), however, no post-vaccination infections were noted with B.1. Post-vaccination infections with delta variant were reported across the age groups (except 0-9 year and 10-19 year, which is explained by the fact that COVID-19 vaccines were not administered to below 18 years age in India until the period of this study) and sex (Fig. 4). Mean age of post-vaccination infection was 44.48(±16.17) year for total cases and 40.4(±11.9) year for men and 47.8 (±20.4) year for women. No significant differences were noted for incidence of post-vaccination infections in terms of age groups and sex.

**Figure 4.**
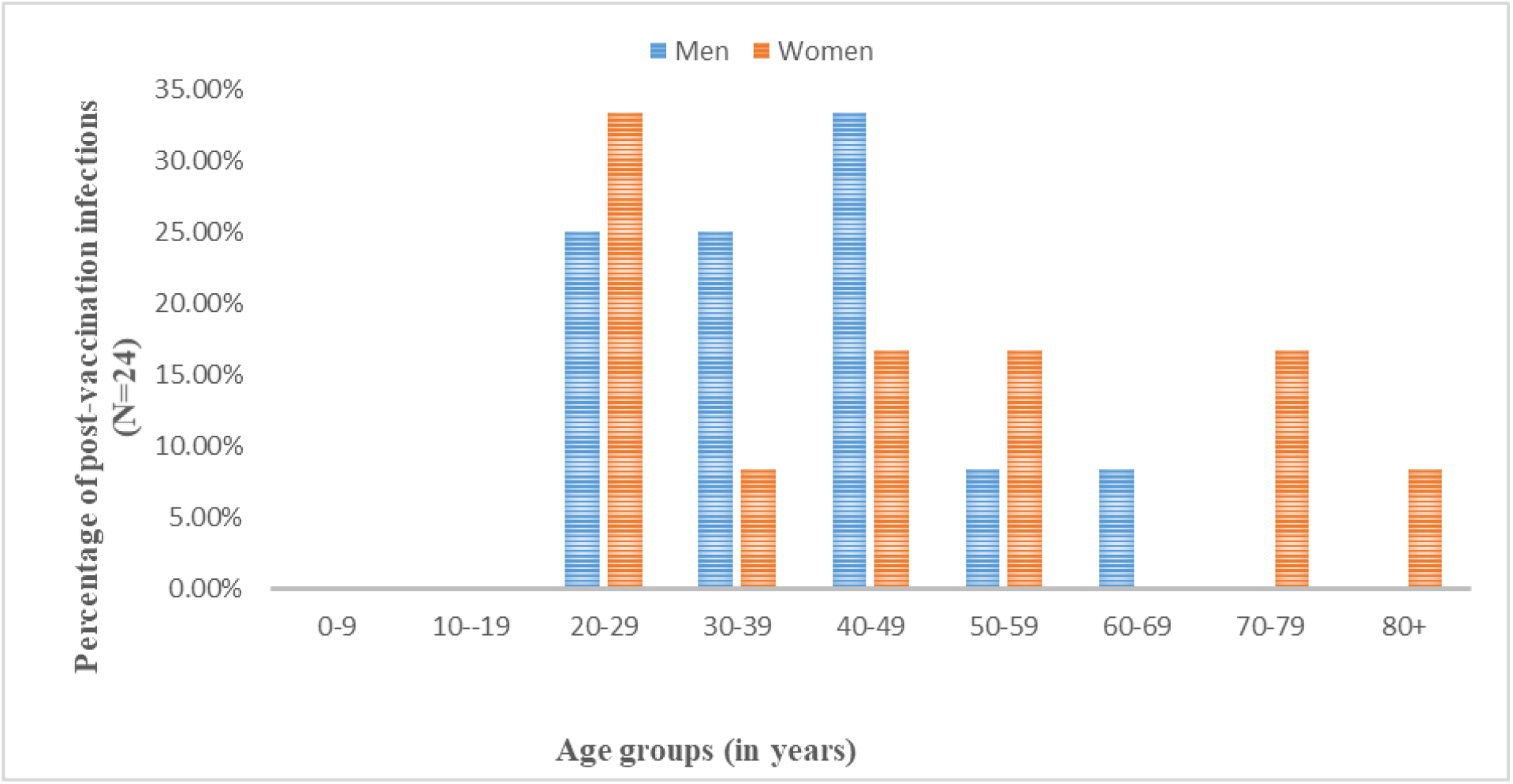
Demographic distribution of post vaccination (two complete doses) SARS-CoV-2 B.1.617.2 (Delta) variant infections. (No post-vaccination infections were noted in 0-9 year and 10-19 year age groups explained by the fact that COVID-19 vaccines were not administered to individuals below 18 year age in India until the period of this study.)

## Discussion

Accumulating evidence indicate that SARS-CoV-2 variants have altered epidemiological characteristics in comparison to WT strains^2,4,22^. Emerging variants are not only more transmissible but are involving younger population in increasing proportion^23–25^, the severity of the illness^25^, and capability for immune escape against natural and acquired immunity^26^ have also been significantly increased in comparison to the WT strains. Although, the delta variant has become globally dominant by now, the studies presenting demographic characteristics for this variant are currently very limited. The B.1 strain was the very first variant with single significant mutation in the spike protein coding region (D614G), and had contributed to the first COVID-19 waves globally. The studies established that it had comparable epidemiological characteristics with original Wuhan strain^4,27^. In this study, we used B.1 strain as a WT control to study the demographic characteristics of delta variant in Indian population. Our analyses unraveled important observations regarding the changing epidemiological characteristics of COVID-19 pandemic (Figs. 1-4, Table 1). In comparison to B.1, with the delta variant higher proportion of young age (<20 year) population contracted infection. The proportion of women contracting infection was also increased. A higher proportion of young (<20 year) population developed symptoms and required hospitalization. Further, sex disproportion was observed in developing symptomatic illness and hospitalization as a higher proportion of young and adult women were affected. The mean age of contracting infection as well as developing symptomatic illness/hospitalization was considerably lower for men as well as women. The total mortality was about 1.8 times higher (13% vs. 7%). Risk of death increased irrespective of the sex (Odds ratio: 3.034, 95% Confidence Interval: 1.7-5.2, p<0.001). Although, the sex of the patients couldn’t predict death, an increase in the proportion of women were noted with delta variant infections. Further, we noted multiple incidences of delta variant infections following two complete doses of the vaccines.

Studies analyzing the demographic involvement by delta variant in Indian population are currently scarce. Recently, G K *et al*^28^ presented their observations in Indian population using data from national clinical registry for COVID-19. The authors studied clinical profiles of the hospitalized patients in the period of second COVID-19 wave, which was mainly driven by delta variant (however, no genomic sequence were informed by the authors to ensure that the analyzed cases only represented those caused by the delta variant). Authors also studied the hospitalized cases from the period of the first COVID-19 wave, which were supposedly caused by WT strains. Authors noted a decrease in the mean age of total population for contracting infection, as well as a reduction in proportion of cases in men during the second wave. Authors further noted that in the second wave mortality among hospitalized patients was 13.26 %, which was by 3.1 percent higher than the first wave. The increase in mortality was seen in all age groups in their study except for <20 year of age, where mortality was decreased^28^. We have observed a similar trend in the susceptibility for contracting infection and risk of mortality in terms of age and sex, except higher increase in total mortality with delta variant in comparison to WT strain was observed in our study (13% vs. 7%), however no deaths were noted in our study in <20 year age groups for both of the strains (Figs. 1-3, Table 1).

India is one of the world’s highest populated country with very high proportion of young (approximately 45 % <20 year)^29^. Our analyses show that, in comparison to B.1, delta variant infections have occurred in considerably higher proportion in <20 year age groups, particularly in 0-9 year age group (4.47% vs. 2.3%) (Fig. 1). The proportion of symptomatic cases and those requiring hospital admission were also increased in young (10% vs. 4%) (Fig. 2a). These findings indicate an age shift for delta variant, as the COVID-19 infections with the WT strains were not common in young, and development of severe symptoms and need of hospitalization was primarily limited to adult, more specifically in elderly^17,18^.

Furthermore, we observed that, although, proportion of men was still higher than women for contracting infection (59% vs. 41%), a higher proportion of women contracted infection (41% vs. 36%) with delta variant. A higher proportion of young (14% vs. 3%) and adult (75% vs. 55%) women than men (7% vs. 4% and 75% vs. 70% for young and adult, respectively) developed symptoms/hospitalized with delta variant in comparison with B.1 (Fig. 2b). Although mortality rate was still higher for men, an increased proportion of women died with delta variant compared to B.1 (32% vs. 25%). Biologically, severity of illness and mortality risks are considered lower for the females in comparison to the males for the infectious diseases for the mammals, including humans^30^. Narrowing the gap between men and women for symptomatic illness and mortality risks with delta variant presents an important epidemiological concern.

Increased involvement of the lower age groups, primarily young, and narrowing the gap for the rate of infection and severity of illness in terms of age and sex in delta variant infections in comparison to the WT strains, have been indicated by the recent studies and surveillance reports from United Kingdom where there has been a surge in delta variant infections^8,31^. Despite of the established immunological privilege to the young^19,20,32^ and women^16^, as were reflected in the infections with WT strains^16,18^, increased risks of contracting infection and severity of illness in these demographic groups, indicates enhanced virulence/lethality in the delta variant.

Lastly, the noted incidences of delta variant infections following two complete doses COVID-19 vaccines in our study (Fig. 4) get supported by multiple recent reports of vaccine breakthrough infections by this variant, including that from the Indian population^9,33^.

Collectively, the findings of this study elaborate on the changing demographic characteristics of COVID-19 pandemic with emergence of delta variant. These findings are important in the aspect, we have only considered those COVID-19 cases for the analysis for which delta variant infections are confirmed by genomic sequencing, ensuring the accuracy of the presented data. These observations can be helpful in deciding health policy measures for preventing recurrent COVID-19 waves as well as in estimating the possible dangers of further variations in delta strain, which are now being reported across the globe^1^.

The findings of this study and others describing demographic involvement in delta variant infections are confirming a shift from the established pattern for the WT strains. There could be multiple factors contributing to this epidemiological shift, most importantly, the variant’s intrinsic properties, such as increased transmissibility, virulence, and immune escape capabilities, as have been indicated by the recent studies^7–9,11,28,34^. A sudden spurt in the number of cases during the second wave and consequent over-burdening of the emergency health response system could have been a possible contributory factor in the increased mortality noted with delta variant. In contrast, the prioritized vaccination of the > 45 year old might have protected vaccinated elderly population and contributed to lowering of the mean age for COVID-19 involvement.

### Limitations of the study

Our study has multiple limitations, which need to be considered while interpreting the findings. Firstly, our data for reporting post-vaccination infections and patient statuses are limited, hence related observations may need further validation from the studies with larger sample size. Secondly, our data for the vaccination status details were not available in most of the accessed genomic sequence reports, hence our data for the post-vaccination infections doesn’t reflect on the prevalence of such cases in the total population. Lastly, we have not taken into account the influence of the confounding factors, such as, comorbidities, vaccination status, and previous COVID-19 infection, etc., on clinical outcomes of the studied cases, which may have influenced the quality of the results.

## Data Availability

Primary data used for this study are publicly available on: SARS-CoV-2 genomic sequence GISAID database: https://www.gisaid.org.The tabulated data can be availed from the corresponding author on reasonable request.

## Acknowledgements

The study has used SARS-CoV-2 genomic sequence data from GISAID (https://www.gisaid.org/), a publically available database.

## Author (s) contributions

AK designed the study and wrote first draft of the manuscript. KR, RKN, RKJ, AS, GK, and PD performed data collection and analysis. AA performed statistical analyses. AA, RKN, CS, CK, MK, RM, GK, HK, KS, SNP, RP, KK, and SK reviewed and edited the manuscript. All authors consented for submission of the final draft.

## Funding declaration

None

## Conflict of Interest

Authors declared no conflicts of interest.

## Ethics statement

An approval from the institute ethics committee was precluded as the data used in this study were retrieved from the publically available database—GISAID (https://www.gisaid.org/).

## Data availability statement

Primary data used for this study are publicly available on: SARS-CoV-2 genomic sequence— GISAID database: https://www.gisaid.org. The tabulated data can be availed from the corresponding author on reasonable request.

## Notes

### Competing Interest Statement

The authors have declared no competing interest.

### Author Declarations

An approval from the institute ethics committee was precluded as the data used in this study were retrieved from the publicly available database Global Initiative on Sharing All Influenza Data (GISAID)(https://www.gisaid.org/).

## References

1. Alaa AL, Julia L. Mullen, MA, et al. B.1.617.2 Lineage Report. http://outbreak.info, (available at https://outbreak.info/situation-reports?pango=B.1.617.2). Accessed 22 September 2021.

2. Khateeb J, Li Y, Zhang H. Emerging SARS-CoV-2 variants of concern and potential intervention approaches. Crit Care 2021 251. 2021;25(1):1–8. doi:10.1186/S13054-021-03662-X

3. McCallum M, Walls AC, Sprouse KR, et al. Molecular basis of immune evasion by the delta and kappa SARS-CoV-2 variants. bioRxiv. Published online August 12, 2021:2021.08.11.455956. doi:10.1101/2021.08.11.455956

4. Kumar A, Parashar R, Faiq MA, et al. Emerging SARS-CoV-2 Variants Can Potentially Break Set Epidemiological Barriers in COVID-19. SSRN Electron J. Published online July 9, 2021. doi:10.2139/SSRN.3888058

5. Planas D, Veyer D, Baidaliuk A, et al. Reduced sensitivity of SARS-CoV-2 variant Delta to antibody neutralization. Nat 2021. Published online July 8, 2021:1-7. doi:10.1038/s41586-021-03777-9

6. Sreelekshmy M, Dhruv YP, Anita S, et al. SARS-CoV-2 Delta variant pathogenesis and host response in Syrian hamsters. bioRxiv. Published online August 4, 2021:2021.07.24.453631. doi:10.1101/2021.07.24.453631

7. Li B, Deng A, Li K, et al. Viral infection and transmission in a large, well-traced outbreak caused by the SARS-CoV-2 Delta variant. medRxiv. Published online July 23, 2021:2021.07.07.21260122. doi:10.1101/2021.07.07.21260122

8. Twohig KA, Nyberg T, Zaidi A, et al. Hospital admission and emergency care attendance risk for SARS-CoV-2 delta (B.1.617.2) compared with alpha (B.1.1.7) variants of concern: a cohort study. Lancet Infect Dis. 0(0). doi:10.1016/S1473-3099(21)00475-8

9. Gupta N, Kaur H, Yadav P, et al. Clinical characterization and Genomic analysis of COVID-19 breakthrough infections during second wave in different states of India. medRxiv. Published online July 15, 2021:2021.07.13.21260273. doi:10.1101/2021.07.13.21260273

10. Campbell F, Archer B, Laurenson-Schafer H, et al. Increased transmissibility and global spread of SARS-CoV-2 variants of concern as at June 2021. Euro Surveill. 2021;26(24). doi:10.2807/1560-7917.ES.2021.26.24.2100509

11. Dhar MS, Marwal R, Ponnusamy K, et al. Genomic characterization and Epidemiology of an emerging SARS-CoV-2 variant in Delhi, India. doi:10.1101/2021.06.02.21258076

12. Yang W, Shaman J. Epidemiological characteristics of three SARS-CoV-2 variants of concern and implications for future COVID-19 pandemic outcomes. medRxiv. Published online May 21, 2021:2021.05.19.21257476. doi:10.1101/2021.05.19.21257476

13. Frampton D, Rampling T, Cross A, et al. Genomic characteristics and clinical effect of the emergent SARS-CoV-2 B.1.1.7 lineage in London, UK: a whole-genome sequencing and hospital-based cohort study. Lancet Infect Dis. 2021;0(0). doi:10.1016/S1473-3099(21)00170-5

14. Faria NR, Mellan TA, Whittaker C, et al. Genomics and epidemiology of the P.1 SARS-CoV-2 lineage in Manaus, Brazil. Science (80-). 2021;372(6544):815–821. doi:10.1126/science.abh2644

15. Funk T, Pharris A, Spiteri G, et al. Characteristics of SARS-CoV-2 variants of concern B.1.1.7, B.1.351 or P.1: data from seven EU/EEA countries, weeks 38/2020 to 10/2021. Eurosurveillance. 2021;26(16):2100348. doi:10.2807/1560-7917.ES.2021.26.16.2100348

16. A K, RK N, M K, et al. COVID-19 pandemic: insights into molecular mechanisms leading to sex-based differences in patient outcomes. Expert Rev Mol Med. 2021;23:e7. doi:10.1017/ERM.2021.9

17. Y C, SL K, BT G, et al. Aging in COVID-19: Vulnerability, immunity and intervention. Ageing Res Rev. 2021;65. doi:10.1016/J.ARR.2020.101205

18. O I, J L, K T, Z W, ZA B. Risk of infection and transmission of SARS-CoV-2 among children and adolescents in households, communities and educational settings: A systematic review and meta-analysis. J Glob Health. 2021;11:1–15. doi:10.7189/JOGH.11.05013

19. Pierce CA, Sy S, Galen B, et al. Natural mucosal barriers and COVID-19 in children. JCI Insight. 2021;6(9). doi:10.1172/JCI.INSIGHT.148694

20. Cohen CA, Li APY, Hachim A, et al. SARS-CoV-2 specific T cell responses are lower in children and increase with age and time after infection. Nat Commun. 2021;12(1). doi:10.1038/S41467-021-24938-4

21. Kumar A, Dwivedi P, Kumar G, et al. Second wave of COVID-19 in India could be predicted with genomic surveillance of SARS-CoV-2 variants coupled with epidemiological data: A tool for future. medRxiv. Published online June 13, 2021:2021.06.09.21258612. doi:10.1101/2021.06.09.21258612

22. Gobeil SM-C, Janowska K, McDowell S, et al. Effect of natural mutations of SARS-CoV-2 on spike structure, conformation, and antigenicity. Science (80-). 2021;373(6555). doi:10.1126/SCIENCE.ABI6226

23. Brookman S, Cook J, Zucherman M, Broughton S, Harman K, Gupta A. Effect of the new SARS-CoV-2 variant B.1.1.7 on children and young people. Lancet Child Adolesc Heal. 2021;5(4):e9–e10. doi:10.1016/S2352-4642(21)00030-4

24. Taylor L. Covid-19: Brazil’s spiralling crisis is increasingly affecting young people. BMJ. 2021;373:n879. doi:10.1136/bmj.n879

25. Nyberg T, Twohig KA, Harris RJ, et al. Risk of hospital admission for patients with SARS-CoV-2 variant B.1.1.7: cohort analysis. BMJ. 2021;373:n1412. doi:10.1136/bmj.n1412

26. Harvey WT, Carabelli AM, Jackson B, et al. SARS-CoV-2 variants, spike mutations and immune escape. Nat Rev Microbiol. 2021;19(7):409–424. doi:10.1038/s41579-021-00573-0

27. Esper FP, Cheng Y-W, Adhikari TM, et al. Genomic Epidemiology of SARS-CoV-2 Infection During the Initial Pandemic Wave and Association With Disease Severity. JAMA Netw Open. 2021;4(4):e217746–e217746. doi:10.1001/JAMANETWORKOPEN.2021.7746

28. G K, A M, RK S, et al. Clinical profile of hospitalized COVID-19 patients in first & second wave of the pandemic: Insights from an Indian registry based observational study. Indian J Med Res. Published online July 14, 2021. doi:10.4103/IJMR.IJMR_1628_21

29. Census of India: Age Structure And Marital Status. Accessed September 7, 2021. https://censusindia.gov.in/census_and_you/age_structure_and_marital_status.aspx

30. Scully EP, Haverfield J, Ursin RL, Tannenbaum C, Klein SL. Considering how biological sex impacts immune responses and COVID-19 outcomes. Nat Rev Immunol. 2020;20(7):442–447. doi:10.1038/s41577-020-0348-8

31. Health England P. SARS-CoV-2 Variants of Concern and Variants under Investigation in England, Technical Briefing 14, 3 June, 2021. https://www.gov.uk/government/publications/investigation-of-novel-sars-cov-2-variant-variant-of-concern-20201201

32. Selva KJ, Sandt CE van de, Lemke MM, et al. Systems serology detects functionally distinct coronavirus antibody features in children and elderly. Nat Commun. 2021;12(1). doi:10.1038/S41467-021-22236-7

33. Hacisuleyman E, Hale C, Saito Y, et al. Vaccine Breakthrough Infections with SARS-CoV-2 Variants. N Engl J Med. 2021;384(23):2212–2218. doi:10.1056/nejmoa2105000

34. Kislaya I, Rodrigues EF, Borges V, et al. Delta variant and mRNA Covid-19 vaccines effectiveness: higher odds of vaccine infection breakthroughs. medRxiv. Published online August 22, 2021:2021.08.14.21262020. doi:10.1101/2021.08.14.21262020

