## Supplementary material for "Demographic characteristics of SARS-CoV-2 B.1.617.2 (Delta) variant infections in Indian population": Fig. S1

### Kumar et al., 2021

Supplementary file 1.

Content: Figure S1.

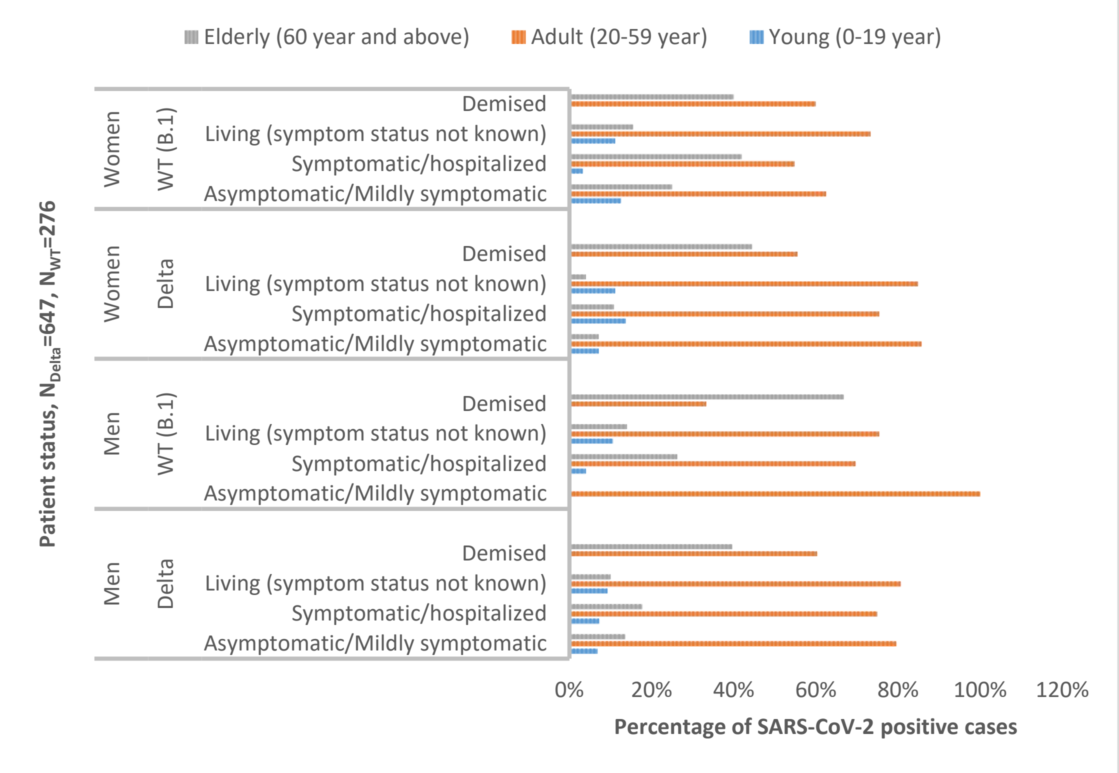


**Figure S1. Distribution of cases with SARS-CoV-2 B.1.617.2 (Delta) variant and Wild type (WT) strain (B.1) infections in reference of patient status.**
